# Wisconsin April 2020 Election Not Associated with Increase in COVID-19 Infection Rates

**DOI:** 10.1101/2020.04.23.20074575

**Authors:** Andrew C. Berry, Madhuri S. Mulekar, Bruce B. Berry

## Abstract

**Background:** Wisconsin (WI) held a primary election in the midst of the COVID-19 pandemic. Live voting at polls was allowed despite concern over increasing the spread of COVID-19. In addition to 1.1 million absentee ballots cast, 453,222 persons voted live. The purpose of our study was to determine if an increase in COVID-19 activity was associated with the election.

**Methods:** Using the voting age population for the United States (US), WI, and its 3 largest counties, and daily new COVID-19 case reports from various COVID-19 web-based dashboards, daily new case rates were calculated. With election day April 7, the incubation period included April 12-21. The new case activity in the rest of the US was compared with the Wisconsin activity during the incubation period.

**Results:** WI daily new case rates were lower than those of the rest of the US for the 10-day period before the election and remained lower during the post exposure incubation period. The ratio of Wisconsin new case rates to US new case rates was 0.34 WI: 1 US for the 10 days leading up to the election and declined to 0.28 WI: 1 US for the 10-day post-incubation period after the election. Similar analysis for Milwaukee county showed a pre-election ratio of 1.02 Milwaukee: 1 US and after the election the ratio was 0.63 Milwaukee: 1 US. Dane county had a pre-election ratio of 0.21 Dane: 1 US case, and it fell to 0.13 Dane: 1 US after the election. Waukesha county had a pre-election ratio of 0.27 Waukesha: 1 US case and that fell to 0.19 Waukesha: 1 US after the election.

**Conclusions:** There was no increase in COVID-19 new case daily rates observed for Wisconsin or its 3 largest counties following the election on April 7, 2020, as compared to the US, during the post-incubation interval period.

## Introduction

Since the World Health Organization declared SARS-CoV-2 (COVID-19) a global pandemic on March 11, 2020, drastic social distancing measures have been declared across the United States^1^and in Wisconsin by Governor Evers Safer at Home Order ^2^. Many businesses and social activities have been closed and minimized, leading to unrest about individual freedoms. The intricate balance between constitutional voting rights and public health took front seat on April 7, 2020, the scheduled primary election date for the state of Wisconsin. The presidential primary election, a key state Supreme Court justice election, and numerous local office elections were on the ballot for April 7.

A U.S. District judge rejected a request to postpone the election, but provided an extension for absentee ballots. Later, the Supreme Court cancelled the extended period for absentee voting,^3^ and on April 7, the election occurred, a mixture of live and absentee mail in voting. Absentee ballots needed to be postmarked by April 7 to count, causing a change in many voter’s plans.

We aimed to see whether a subsequent rise in COVID-19 cases followed the controversial in-person Wisconsin election on April 7, 2020. Not only may this impact local public health actions, but could impact future election behaviors.

## Methods

Websites for new COVID-19 daily cases were visited daily to obtain new case data for the United States (US)^4^, Wisconsin^5^, Milwaukee county^6^, Dane county^7^, and Waukesha county^8^. Those counties represented the largest 3 contingents of voter age adults in Wisconsin and were counties where COVID-19 was active during election time. The number of new cases reported by the websites daily were extracted, and the daily new case rates per 100,000 voting age (age 18 or older) adults were calculated in those populations. Census data^9^ was used to obtain population data and age mix, then the number of voter age adults were calculated for the cohorts. The number of Wisconsin (WI) voters was subtracted from the US total, so the US data would represent all of the country excluding WI data. Daily new cases of COVID-19 infections extracted from the websites were then divided by the number of voter age adults in the cohorts to determine the daily new case rates. COVID-19 daily cases numbers from WI were removed to determine the number of new daily cases for US. The daily new COVID-19 cases were not adjusted for age (<3% under age 19)^5^. The study was exempt from IRB, it did not include protective health information and used data in the public domain exclusively.

The median incubation period of the virus is 5 days^10^. With the election on April 7, we used April 12 as the first date to start monitoring the number of new COVID-19 cases that may be related to the election. We then continued the analysis for the full 14-day period following exposure, mimicking a self-quarantine period, as <1% are shown to develop symptoms after 14 days^10^. Analysis of data from April 12-21 best represents the viral properties and course of action by the individual, from symptoms to testing to receiving results to being reported by the local health department.

## Results

Wisconsin’s April 7, 2020 election was completed and allowed live in-person voting, with subsequent voting characteristics listed in Table 1, for the three largest voting counties, state of Wisconsin, and US, which show considerably large number of live in-person voters.

**Table 1:**
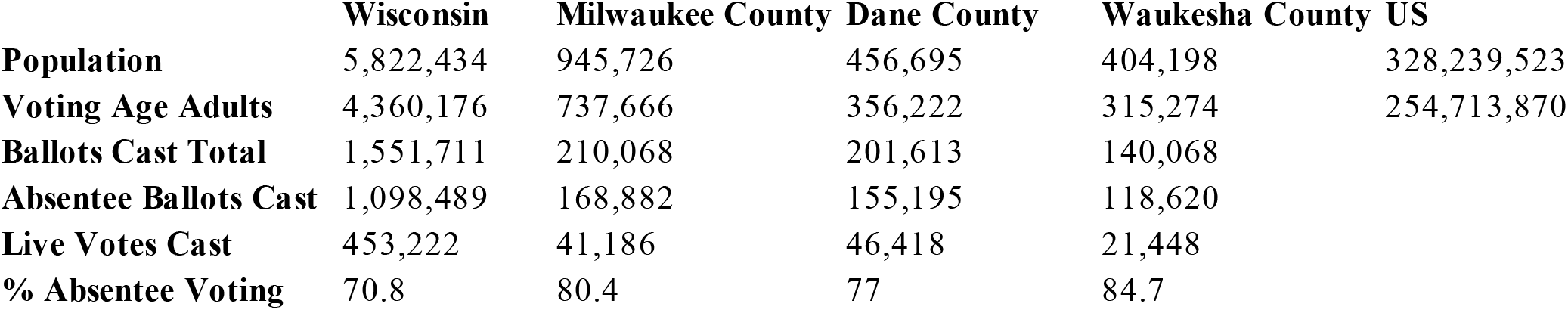
Election Demographics for Wisconsin, Wisconsin Three Major Counties, and United States (US)

Figure 1 displays the daily rate of new cases of COVID-19 by day of the pandemic, for Wisconsin and the rest of the US, with curves visually mimicking each other. Figure 1 displays election day occurring on day 28 (highlighted), and the incubation period occurred during days 33-42, five days post-election. Figure 2 provides a focused comparison between Wisconsin and the US for the 5-14 days post-election period. It does not suggest any spike in post-election cases in Wisconsin in relation to the rest of the US.

**Figure 1:**
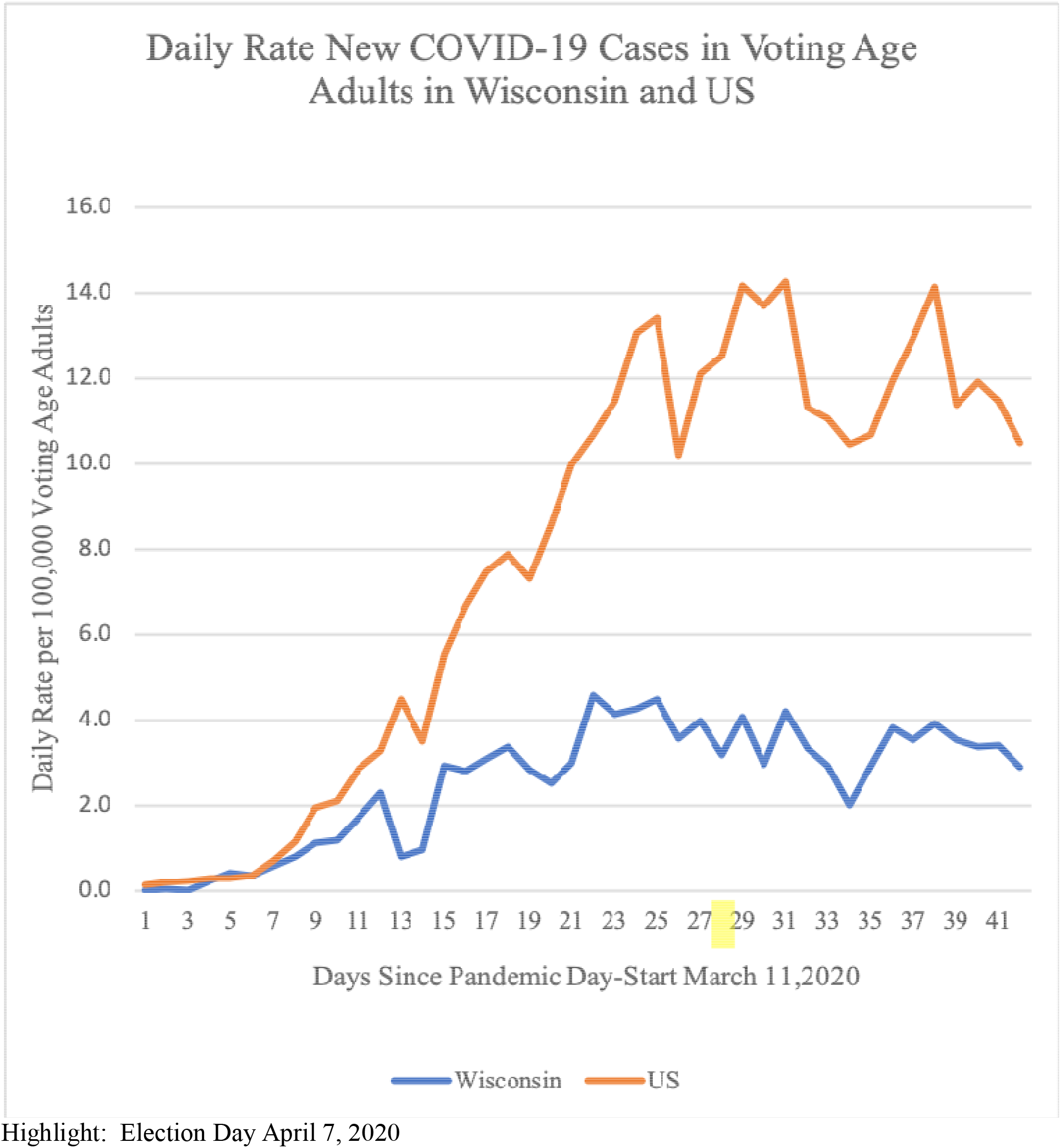

**Figure 2:**
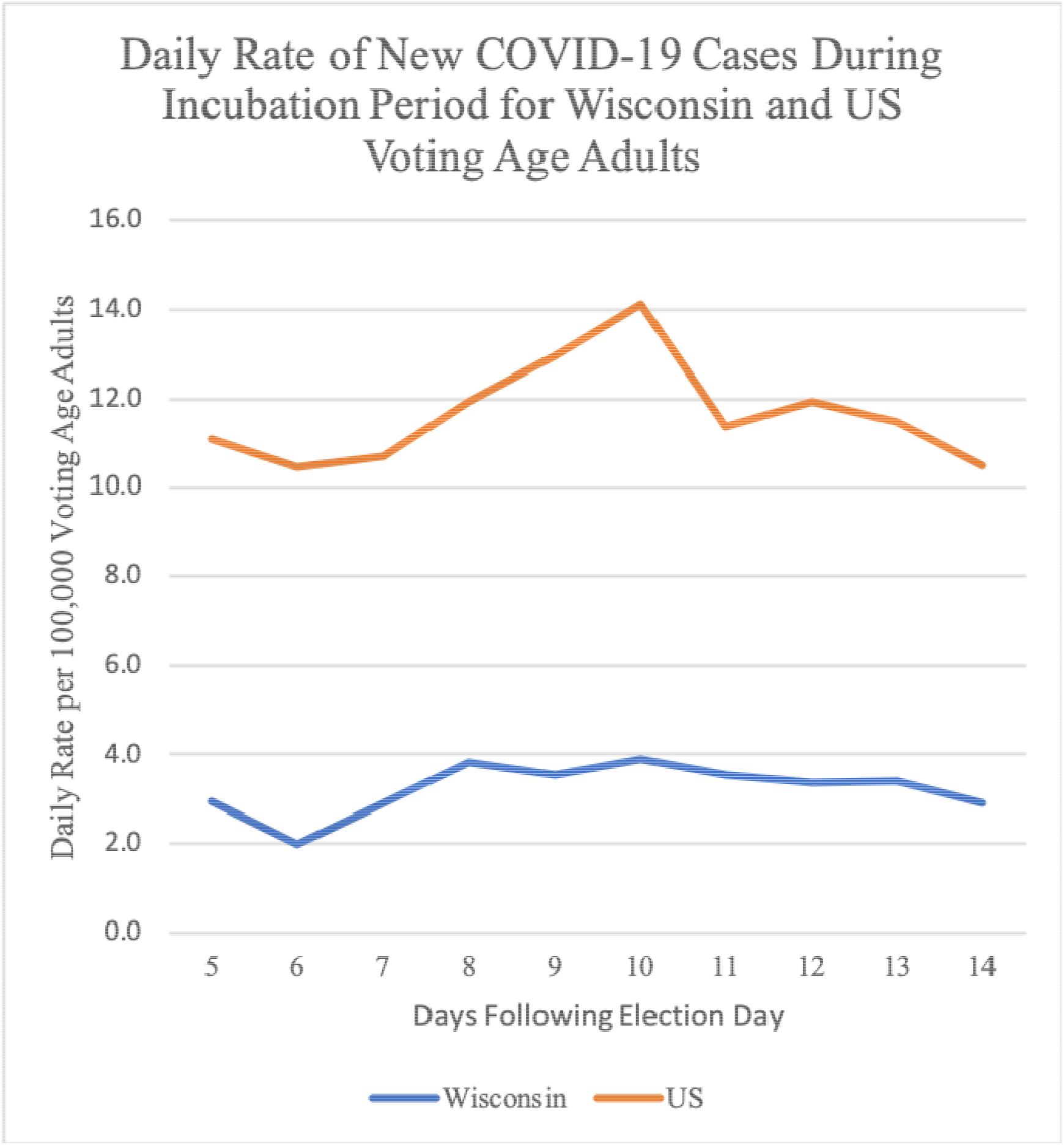

The average rate of new cases of COVID-19 was 10.77 per 100,000 voting age adults (all rates are per 100,000 voting age adults) in the US for the 10 days leading up to the election on April 7, 2020, and 11.62 for the 10-day incubation period following the election (April 12-21) (Table 2, Supplement A-B). Using the same time period, the average daily rate of new COVID-19 cases for Wisconsin was 3.65 before the election, and 3.23 for the 10-day incubation period following the election.

**Table 2:** Pre and Post-Election Daily New Case Rates and Ratios Comparing United States (US), Wisconsin, and Wisconsin Three Major Counties.

|  | Mean Daily Rate New Cases per 100,000 Voting Age Adults (with Ratios) |  |  |  |  |  |  |  |  |
| --- | --- | --- | --- | --- | --- | --- | --- | --- | --- |
|  | US | WI | WI:US | Milwaukee | Milwaukee:US | Dane | Dane:US | Waukesha | Waukesha:US |
| **PRE-Election 10-Day Period** | 10.77 | 3.65 | 0.34 | 10.97 | 1.02 | 3.68 | 0.21 | 2.92 | 0.27 |
| **POST-Election 10-Day Period** | 11.62 | 3.23 | 0.28 | 7.28 | 0.63 | 1.54 | 0.13 | 2.16 | 0.19 |
| (Days 5-14 Post Election) |  |  |  |  |  |  |  |  |  |

The US and WI rates themselves, are of course different as the circumstances in the US are different than those in Wisconsin as it pertains to the course of pandemic, population mix, population densities, and many other factors. So, a ratio was determined to see if the correlation between the US and Wisconsin was consistent. Prior to the election, the Wisconsin daily rate of new COVID-19 cases compared to that for the US rate was in a ratio of 0.34 WI:1 US (Table 2, Supplement A-B). The average daily new case rate after the election was 11.62 in the US compared to 3.23 in Wisconsin, with a ratio of 0.28 WI:1 US (Table 2, Supplement A-B). After the election the ratio of new daily case activity in Wisconsin compared to the rest of the US dropped from its pre-election level, suggesting the rate of development of new cases was decreasing following the election compared to what would have been expected if the relationship between Wisconsin and the rest of the US had continued at its pre-election ratio.

The daily rate of new cases of COVID-19 by day of the pandemic for each of the three Wisconsin counties with the most voting age residents—Milwaukee, Dane, and Waukesha—are shown in Figure 3, Figure 4, and Figure 5, respectively. The figures do not suggest any significant spike in cases in any one of these three counties as compared to the US during that time. Milwaukee county’s average rate of daily new COVID-19 cases (10.97) was nearly the same as the US (10.77), but higher than the Wisconsin rate of 3.65 for the 10 days prior to the election (Table 2, Supplement A-B). Following the election, the Milwaukee county rate dropped to 7.28, while the US rate increased to 11.62. Prior to the election the ratio between Milwaukee county and the US was 1.02 Milwaukee: 1 US. After the election the ratio went down to 0.63 Milwaukee: 1 US, which is consistent with a drop in rate of new cases in Milwaukee county beyond what would have been expected should the relationship between Milwaukee county data and the rest of the US had continued at the pre-election ratios.

**Figure 3:**
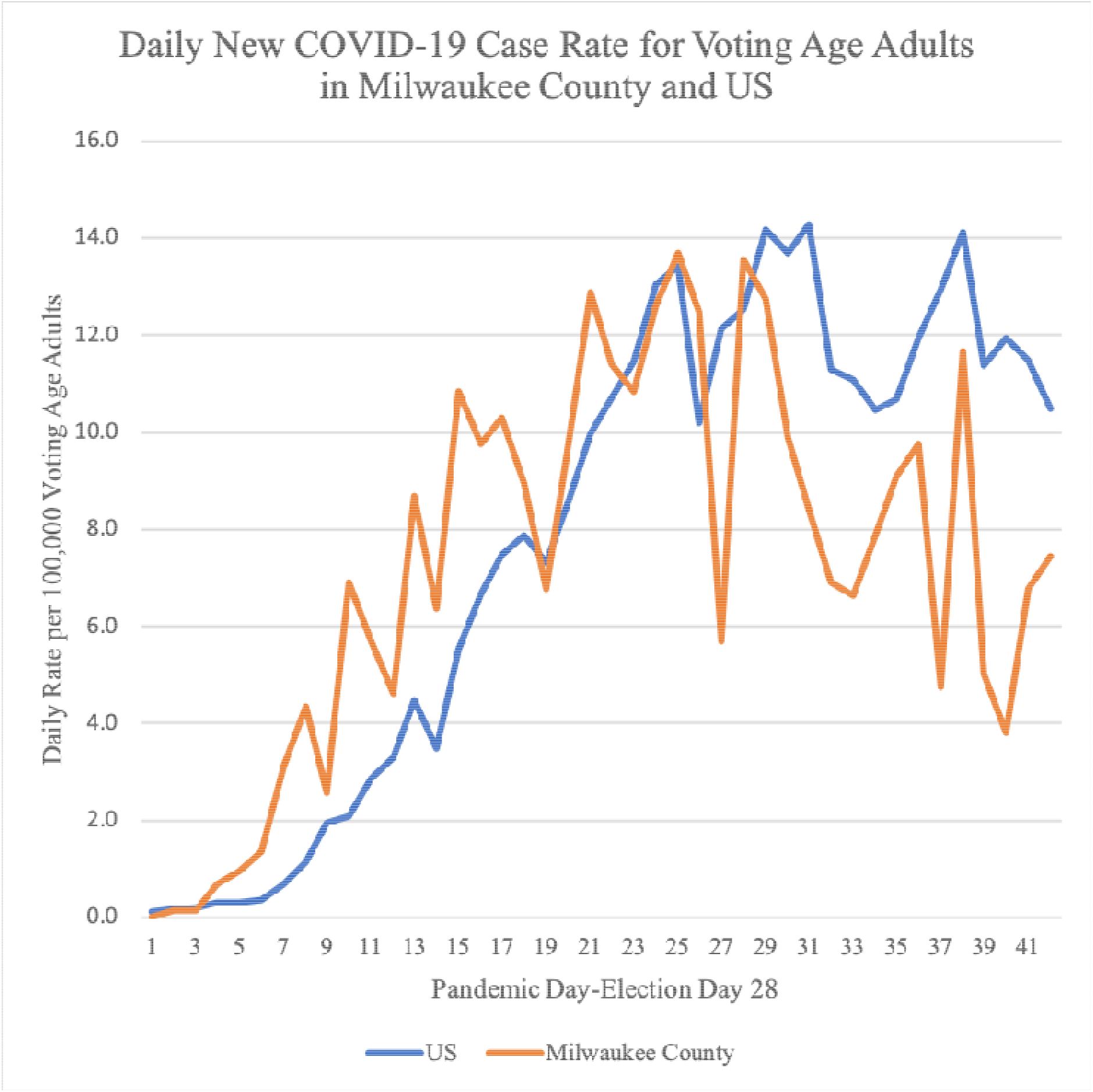

**Figure 4:**
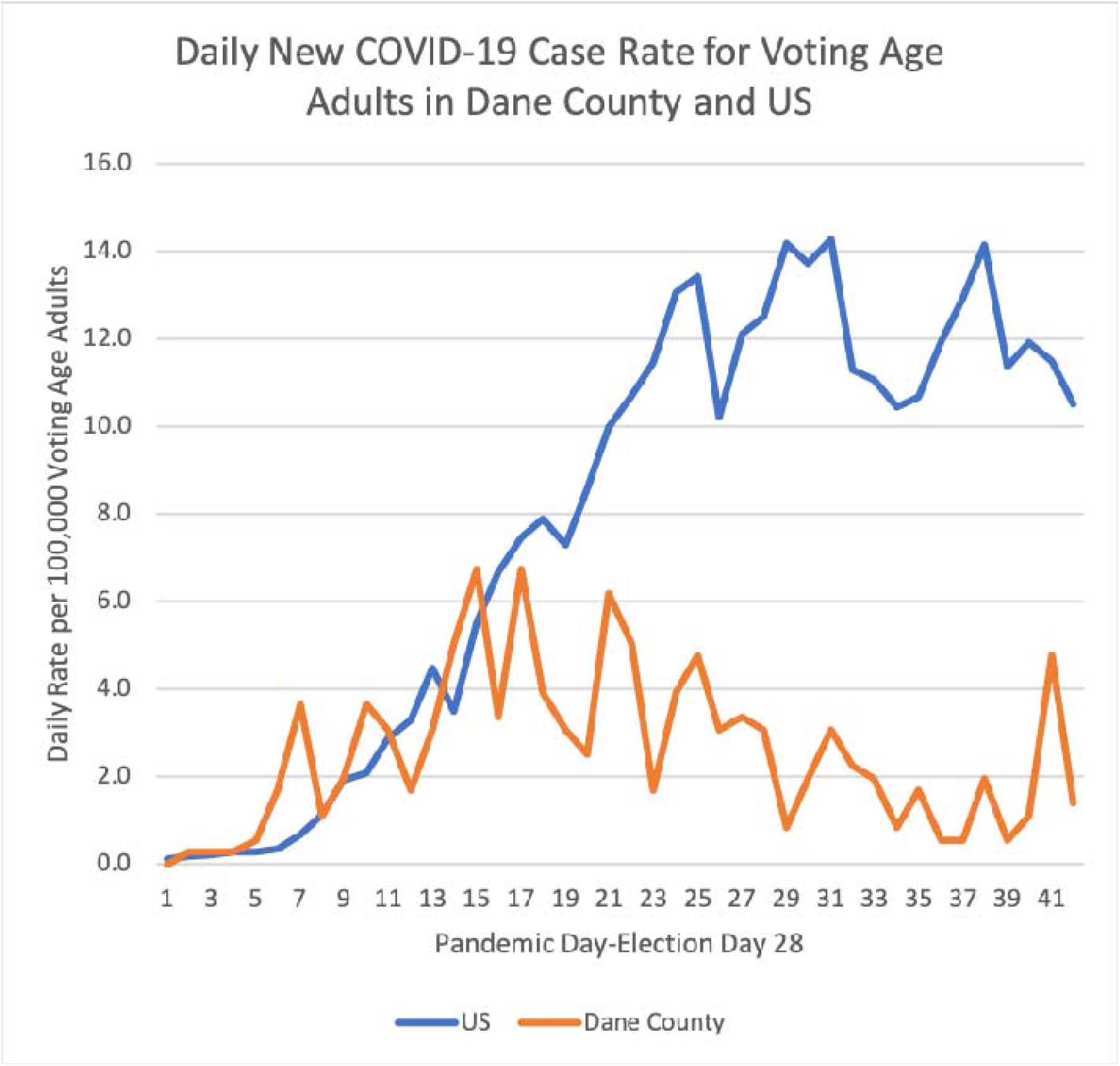

**Figure 5:**
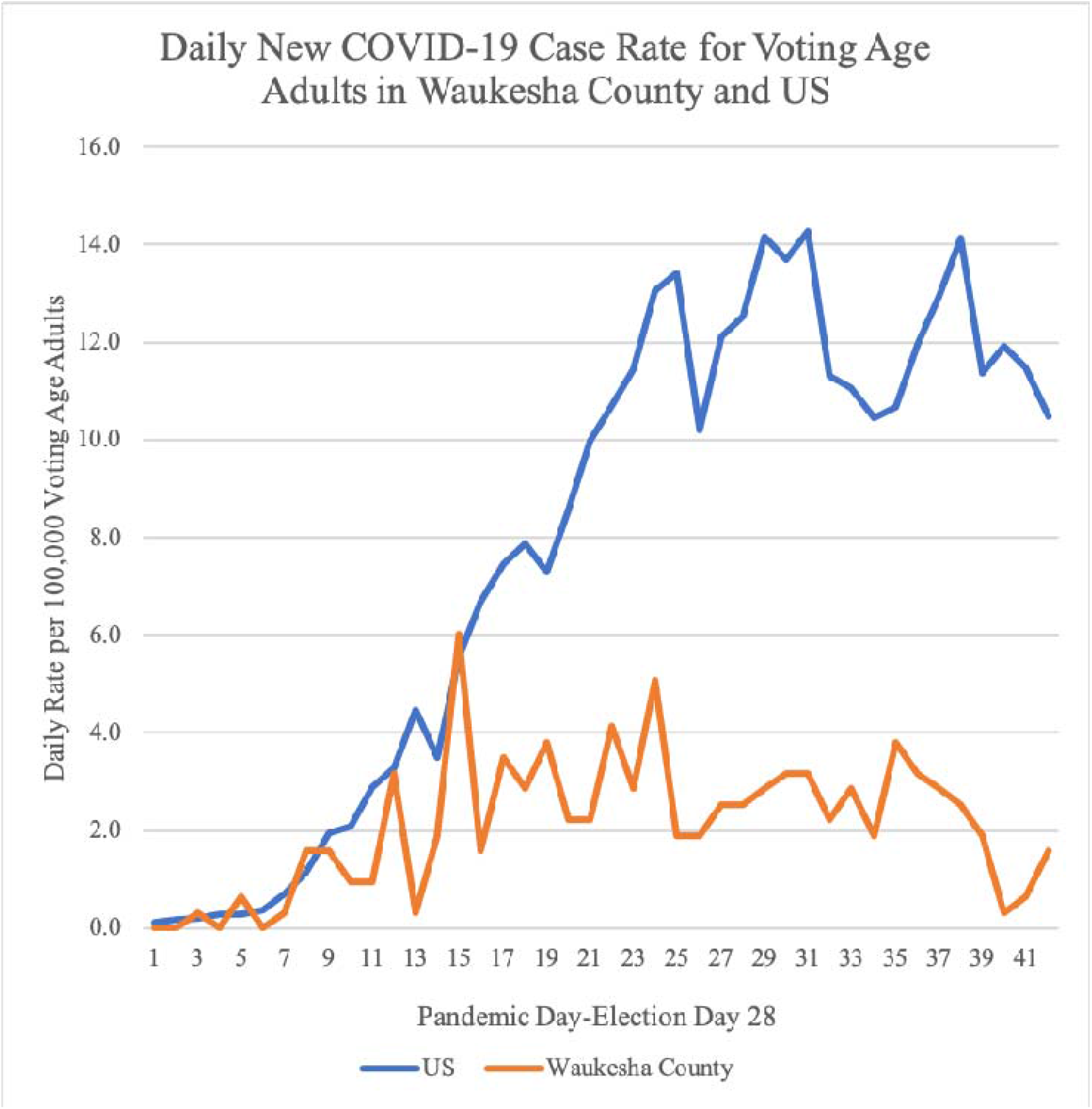

A reduction in post-election new case rates was found for Dane county as well. Dane county started with an average new case daily rate of 3.68 which was lower than the US average (10.77) and it further dropped to 1.54 following the election. The ratio of new cases between Dane and the US was 0.21 Dane: 1 US before the election and 0.13 Dane: 1 US after the election (Table 2, Supplement A-B). Similarly, Waukesha county started with an average new daily case rate of 2.92 prior to the election and it dropped to 2.16 after the election. The ratio of new case rates between Waukesha county and the rest of the US was 0.27 Waukesha: 1 US prior to the election and 0.19 Waukesha: 1 US after the election (Table 2, Supplement A-B). Both Dane and Waukesha counties showed a drop in new cases rates beyond what would have been expected should the relationship between the counties’ new case rates and the rest of the US had continued at the pre-election ratios.

## Discussion

Our study did not find any significant increase in the rate of new COVID-19 cases following the April 7, 2020 election post-incubation period, for the state of Wisconsin or its three major voting counties, as compared to the US. Ethically, it is not possible to design a randomized study to investigate associations between an in-person voting event and the development of new COVID-19 symptoms. The next best option would be to study two groups of people, matched for all the known risks factors for contracting COVID-19 such as age, gender, race, diabetes, hypertension, occupation, sick contacts, etc—but this remains an arduous task that is impractical. Our study compared the rate of new COVID-19 cases following the Wisconsin election to the rest of the US. Thus, we took a practical approach and observed that the COVID-19 activity in Wisconsin seemed to parallel the activity in the rest of the United States (Figure 1, Figure 2). Prior studies have shown that most people who are going to show symptoms do so between 5-14 days following an exposure^9^. Thus, a 10-day period before the election was used to establish a ratio reflecting the relationship between Wisconsin and US rates and then was compared with the ratio observed during the 10-day incubation period following the potential COVID-19 exposure during the in-person election. A reduction in daily new case rates in Wisconsin was observed compared to what would have been expected if the rates in Wisconsin had followed the pre-election ratios. Our initial hypothesis of an increase in COVID-19 activity following the live election was not supported. There is no scientific reason why the election would cause a reduction in COVID-19 cases and there is nothing about voting that seems protective against COVID-19. The explanation may lie in characteristics and behaviors of those involved.

The concern that live voting in Wisconsin would cause a large spike in COVID-19 cases caused considerable turmoil in the days prior to election and an increase in absentee voting, that may have been a large factor in preventing an increase in COVID-19 activity. There were 1,551,711 absentee ballots cast, but there were 453,222 ballots^11^ cast by voters who went to polls to vote and many stood in line for hours. With the heightened publicity around COVID-19 and the perceived risks associated with voting live, high-risk individuals may have self-selected themselves out of the live voting process. Protective measures at the polls may also have mitigated some of the risk associated with the increased social exposure. Maybe the characteristics of the live voters were more favorable to producing asymptomatic infections and many went undetected. A mixture of all those things likely contributed to the absence of an increase in daily new case daily rates following the election. In the three most populous Wisconsin counties, where 109,052 live ballots were cast, no significant increase in daily rates compared to the rest of the US (Figures 3-5) was observed. Similar to Wisconsin as a whole, those counties’ daily new case rates fell faster compared to the rest of the US as well.

Individual cases of COVID-19 infections most likely occurred as a result of additional exposure from live voting. Contact tracing is very difficult at this time with potential for exposure virtually everywhere in the community, linking an individual to live voting as their sole risk is not possible. Although a designed experiment is not possible to answer the questions raised, the next best option would be a retrospective study where two groups of people are matched for all the known risks factors for contracting COVID-19 such as age, gender, race, diabetes, hypertension and many others. In a future retrospective study, one could extract characteristics about those people who voted live and those voted absentee, and then compare the new COVID-19 case rates for those with comparable characteristics. Even if the two groups could be matched for many of the known risk factors associated with developing COVID-19, it will still not be easy to control for their other exposures, activities of daily living, other household members, and the risk factors and behaviors of other household members.

## Conclusions

No evidence was found to support an increase in COVID-19 new daily case rates for the state of Wisconsin, nor its major voting counties, compared to the rest of the US following live voting on April 7,2020. We must continue to utilize our knowledge about COVID-19 and social distancing measures to create the safest and most effective voting environment for all.

## Data Availability

All data is provided in the manuscript and supplement tables.

## Funding

No funding by any organization or political member for this project or to any member of the research team.

## Contributions

ACB and BBB both conceptualized and designed the study. MSM conducted data analysis. ACB, BBB, MSM designed, prepared, and finalized the manuscript. BBB is the article guarantor.

## Competing Interest

ACB, BBB, MSM have nothing to declare.

---

**Supplement Table A:**
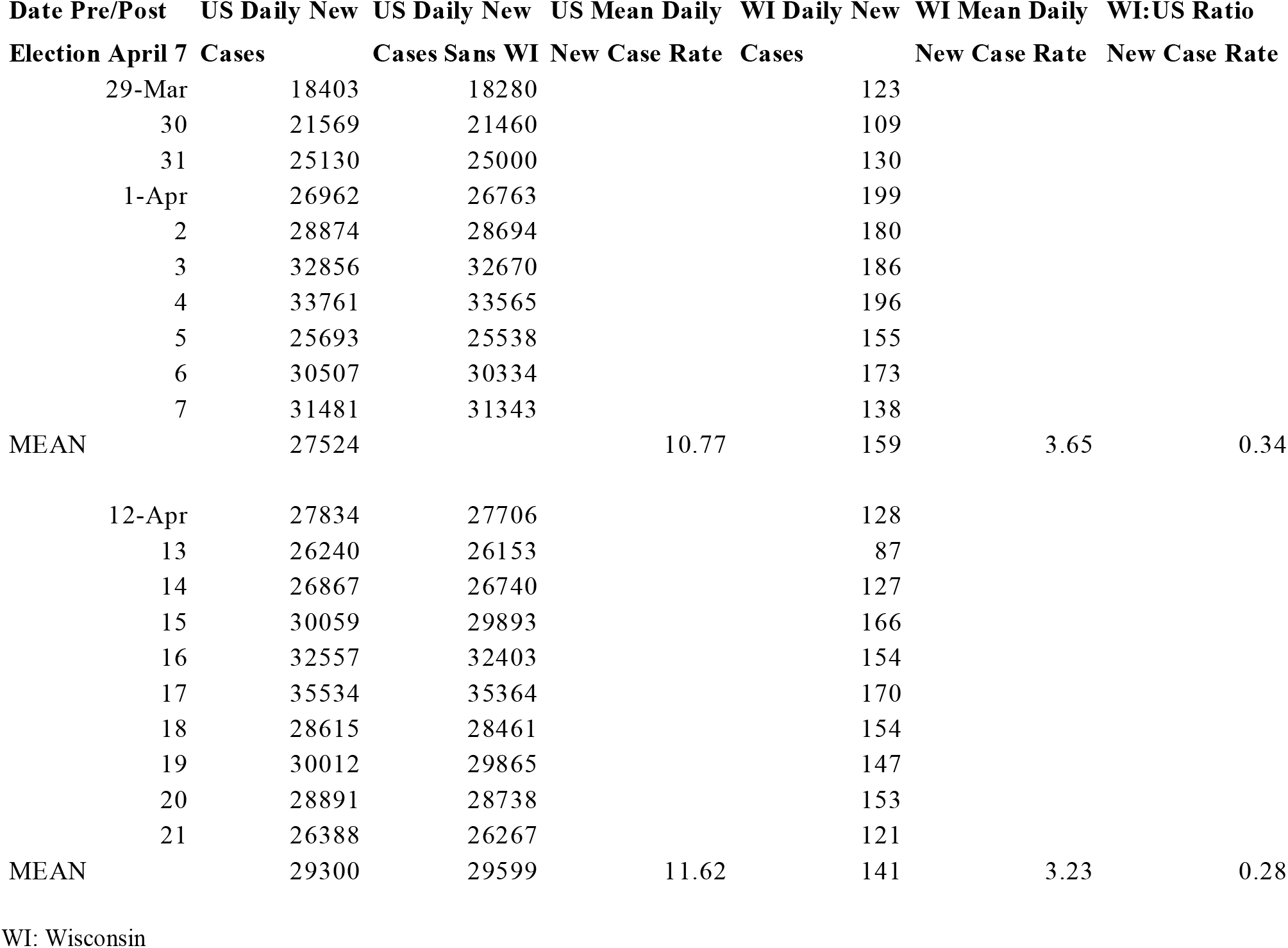
Complete Data of Pre and Post-Election Daily New Case Rates and Ratios Comparing United States (US) and Wisconsin.

**Supplement Table B:** Complete Data of Pre and Post-Election Daily New Case Rates and Ratios Comparing United States (US) and Three Major Wisconsin Counties.

| Date Pre/Post Election | MKE Daily New Cases | MKE Mean Daily New Case Rate | MKE:US Ratio New Case Rate | DANE Daily New Cases | DANE Mean Daily New Case Rate | DANE:US Ratio New Case Rate | WAUK Daily New Cases | WAUK Mean Daily New Case Rate | WAUK:US Ratio New Case Rate |
| --- | --- | --- | --- | --- | --- | --- | --- | --- | --- |
| 29-Mar | 50 |  |  | 11 |  |  | 12 |  |  |
| 30 | 72 |  |  | 9 |  |  | 7 |  |  |
| 31 | 95 |  |  | 22 |  |  | 7 |  |  |
| 1-Apr | 84 |  |  | 18 |  |  | 13 |  |  |
| 2 | 80 |  |  | 6 |  |  | 9 |  |  |
| 3 | 93 |  |  | 14 |  |  | 16 |  |  |
| 4 | 101 |  |  | 17 |  |  | 6 |  |  |
| 5 | 92 |  |  | 11 |  |  | 6 |  |  |
| 6 | 42 |  |  | 12 |  |  | 8 |  |  |
| 7 | 100 |  |  | 11 |  |  | 8 |  |  |
| MEAN | 80.9 | 10.97 | 1.02 | 13.1 | 3.68 | 0.21 | 9.2 | 2.92 | 0.27 |
| 12 | 49 |  |  | 7 |  |  | 9 |  |  |
| 13 | 58 |  |  | 3 |  |  | 6 |  |  |
| 14 | 67 |  |  | 6 |  |  | 12 |  |  |
| 15 | 72 |  |  | 2 |  |  | 10 |  |  |
| 16 | 35 |  |  | 2 |  |  | 9 |  |  |
| 17 | 86 |  |  | 7 |  |  | 8 |  |  |
| 18 | 37 |  |  | 2 |  |  | 6 |  |  |
| 19 | 28 |  |  | 4 |  |  | 1 |  |  |
| 20 | 50 |  |  | 17 |  |  | 2 |  |  |
| 21 | 55 |  |  | 5 |  |  | 5 |  |  |
| MEAN | 53.7 | 7.28 | 0.63 | 5.5 | 1.54 | 0.13 | 6.8 | 2.16 | 0.19 |

MKE: Milwaukee WAUK: Waukesha

## Notes

### Competing Interest Statement

The authors have declared no competing interest.

### Clinical Trial

n/a

### Funding Statement

None

